# Characterizing preserved autonomic regulation following spinal cord injury: Methods of a novel concerted testing battery and illustrative examples of a new translationally focused data representation

**DOI:** 10.1101/2024.05.31.24308290

**Authors:** Ryan Solinsky, Kathryn Burns, Jason W. Hamner, Wolfgang Singer, J. Andrew Taylor

## Abstract

Autonomic dysfunction is common after spinal cord injury, though differing from motor and sensory function, there are currently no established batteries of tests to comprehensively characterize these deficits. Further, while individual established autonomic tests have a long history and sound scientific background, translating these autonomic testing results to inform clinical understanding is a major barrier. Herein, we outline a battery of six laboratory autonomic tests which were carefully curated to collectively describe the ability of individuals with spinal cord injury to inhibit and recruit sympathetic activity through the injured spinal cord. Presenting normative control data in 23 uninjured individuals completing this testing battery, we further demonstrate the utility of extracting three key testing metrics for each test, comparing these control results to 11 individuals with spinal cord injury. Results demonstrate strong normality of data with testing psychometrics suggesting reliable reproducibility on repeat testing. Further, even in this preliminary sample of individuals with spinal cord injuries, clear differences begin to emerge. This illustrates the ability of this collective testing battery to characterize autonomic regulation after spinal cord injury. To aid in clinical translation, we further present a graphical representation, an *autonomic phenotype*, which serves as a snapshot of how normal or abnormal sympathetic inhibition and recruitment of activation may be after spinal cord injury. Utilizing these *autonomic phenotypes*, three example cases of individuals with spinal cord injury highlight evidence of varied degrees of autonomically complete spinal cord injury. Together, this represents a key advancement in our understanding of autonomic function after spinal cord injury.

## Introduction

Though paralysis is the most visible manifestation of spinal cord injury (SCI), secondary autonomic complications such as autonomic dysreflexia and orthostatic hypotension are extremely common.^1^ Broadly, secondary autonomic complications degrade quality of life, utilize significant healthcare resources, and routinely rank as top concerns of individuals with SCI.^2-4^ Despite the clear regulatory role of the autonomic nervous system in healthy physiology, after SCI, there has been only limited characterizations of resultant autonomic dysfunction.^5,6^

The autonomic nervous system regulates the heart and vasculature, skin blood flow, and sweating functions, all systems where dysregulation is a hallmark of SCI.^7^ Cardiovascular autonomic dysfunction after SCI has significant clinical repercussions. Autonomic dysreflexia can be an acute, emergent condition, but can also contribute to the disproportionately high risk of cardiovascular disease in these patients.^8^ In autonomic dysreflexia, an unperceived noxious stimulus such as bladder or bowel distension^9^ below the neurological level of injury induces a sympathetic cascade, causing vasoconstriction below the level of injury and resultant increases in blood pressure that the body must attempt to buffer through the baroreflex.^10,11^ Depending on the degree of preserved sympathetic control through the injury and secondary anatomic changes in the spinal cord, individuals’ ability to buffer these changes can vary widely. However, these altered physiologic interactions may quickly multiply in complexity. Mild tachycardia is common in SCI with autonomic dysreflexia.^12^ This may result from decreased baroreflex sensitivity,^13^ increased arterial stiffness,^14^ or even incomplete sensory lesions leading to partially intact descending supralesional sympathetic activity. In orthostatic hypotension, inability to activate sympathetic vasoconstriction below the level of injury causes blood pressure to fall precipitously with postural change.^15^ Clinically, this may lead to cognitive decline or loss of consciousness. As risk of both autonomic dysreflexia and orthostatic hypotension commonly coexist within individuals with SCI, firmly understanding this underlying autonomic physiology is critical.

Characterizing cardiovascular autonomic deficits is possible with strategically designed testing protocols, as has been done for a number of other diseases.^16-18^ Laboratory-based cardiovascular autonomic measurements have existed for over a century,^19,20^ establishing a robust toolset, though one which has rarely been applied in a systematic way to individuals with SCI. As SCI is a unique condition with often discrete disconnections within the autonomic nervous system, manifesting as above level regions of normal control and below level regions of variably unregulated and amplified self-governance, a strategic testing approach created with this in mind is necessary. Our goals in this study have been twofold: 1) Test a comprehensive battery of autonomic tests to characterize autonomic regulation specifically targeted for individuals with SCI; 2) Develop a novel graphical representation of this data to facilitate understanding of specific autonomic dysfunction after SCI.

## Methods

Following approval by our local IRB, both individuals with SCI and healthy, uninjured controls were prospectively enrolled (NCT04493372). Individuals with SCI were included if they were 18-50 years old and had a history of traumatic SCI, in addition to a matched, uninjured cohort to serve as controls. In an effort to maximize representation, American Spinal Injury Association Impairment Scale A-D injuries were included, with neurological levels of injury from C1-T12.^21^ Exclusion criteria were a history of clinically diagnosed cardiovascular disease, hypertension, diabetes, current pregnancy, or neurological disorder with exception of SCI. Uninjured controls with body mass index of greater than 30 kg/m^2^ were excluded to limit early, potentially undiagnosed, cardiovascular disease and diabetes in the control cohort (Table 1).

**Table 1:**
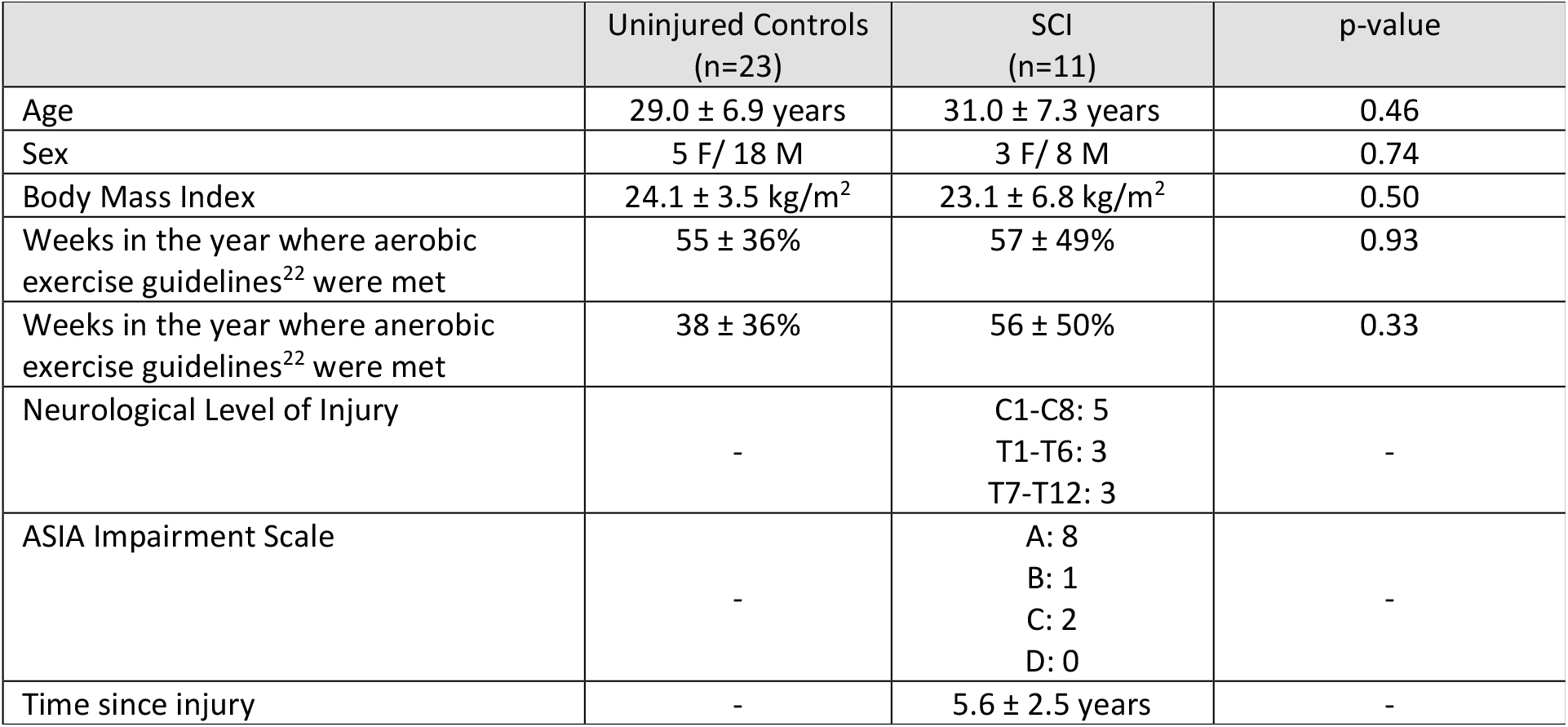
Baseline demographics for uninjured controls. Data presented as mean ± standard deviation. F= female; M= male

|  | Uninjured Controls<br>(n=23) | SCI<br>(n=11) | p-value |
| --- | --- | --- | --- |
| Age | 29.0 ± 6.9 years | 31.0 ± 7.3 years | 0.46 |
| Sex | 5 F/ 18 M | 3 F/ 8 M | 0.74 |
| Body Mass Index | 24.1 ± 3.5 kg/m<sup>2</sup> | 23.1 ± 6.8 kg/m<sup>2</sup> | 0.50 |
| Weeks in the year where aerobic exercise guidelines<sup>22</sup> were met | 55 ± 36% | 57 ± 49% | 0.93 |
| Weeks in the year where anerobic exercise guidelines<sup>22</sup> were met | 38 ± 36% | 56 ± 50% | 0.33 |
| Neurological Level of Injury | - | C1-C8: 5<br>T1-T6: 3<br>T7-T12: 3 | - |
| ASIA Impairment Scale | - | A: 8<br>B: 1<br>C: 2<br>D: 0 | - |
| Time since injury | - | 5.6 ± 2.5 years | - |

To characterize autonomic regulation after SCI, a battery of laboratory tests was established to specifically identify the ability of the autonomic nervous system to inhibit sympathetic activity, recruit sympathetic activity from above the level of injury, and recruit sympathetic activity from below the level of injury.

### Laboratory Testing Paradigm

All participants presented to our autonomic laboratory between 7:30 and 10:00 AM to minimize circadian fluctuations in baseline sympathetic activity.^23^ All had abstained from caffeine and exercise for at least 12 hours and had either fasted that morning or eaten only a small meal greater than 3 hours before testing.^24^ Following consent, all individuals completed a health history as well as the Autonomic Dysfunction Following SCI (ADFSCI) questionnaire^25^ and the Composite Autonomic Symptom Score (COMPASS-31).^26^ The ADFSCI specifically quantifies symptomatic blood pressure regulation problems individuals with SCI may be having as a result of autonomic dysfunction. The COMPASS-31 is a validated scoring metric to broadly quantify autonomic symptoms across six domains: orthostatic intolerance, vasomotor, secretomotor, pupillomotor, bladder, and gastrointestinal. All individuals were also asked about their exercise habits as it pertained to meeting SCI-specific guidelines for aerobic and anerobic activity.^22^ Individuals with SCI further underwent formal neurological evaluation of their injury with the International Standards for Neurological Classification of Spinal Cord Injury exam by a certified physician examiner.^21^

All individuals then emptied their bladders to minimize noxious stimuli during testing and were instrumented for continuous physiologic recordings (Figure 1). Heart rate was recorded with a 5-lead EKG (Dinamap Dash 2000, GE) and beat-to-beat blood pressure was obtained using a finapres finger cuff (AD Instruments), calibrated throughout testing with an automated brachial cuff (Dinamap Dash 2000, GE) between each of the six component tests. Respiratory excursion (Respitrace, SensorMedics) and lower limb blood flow with 4-MHz popliteal ultrasound (Doppler-BoxX, DWL) were recorded. Utilizing blood pressure and flow recordings, lower extremity vascular resistance was calculated. With instrumentation in place, participants were given 15 minutes of supine rest to equilibrate in our temperature-controlled lab (21° C). Participants then underwent the following laboratory-based testing battery.

**Figure 1:**
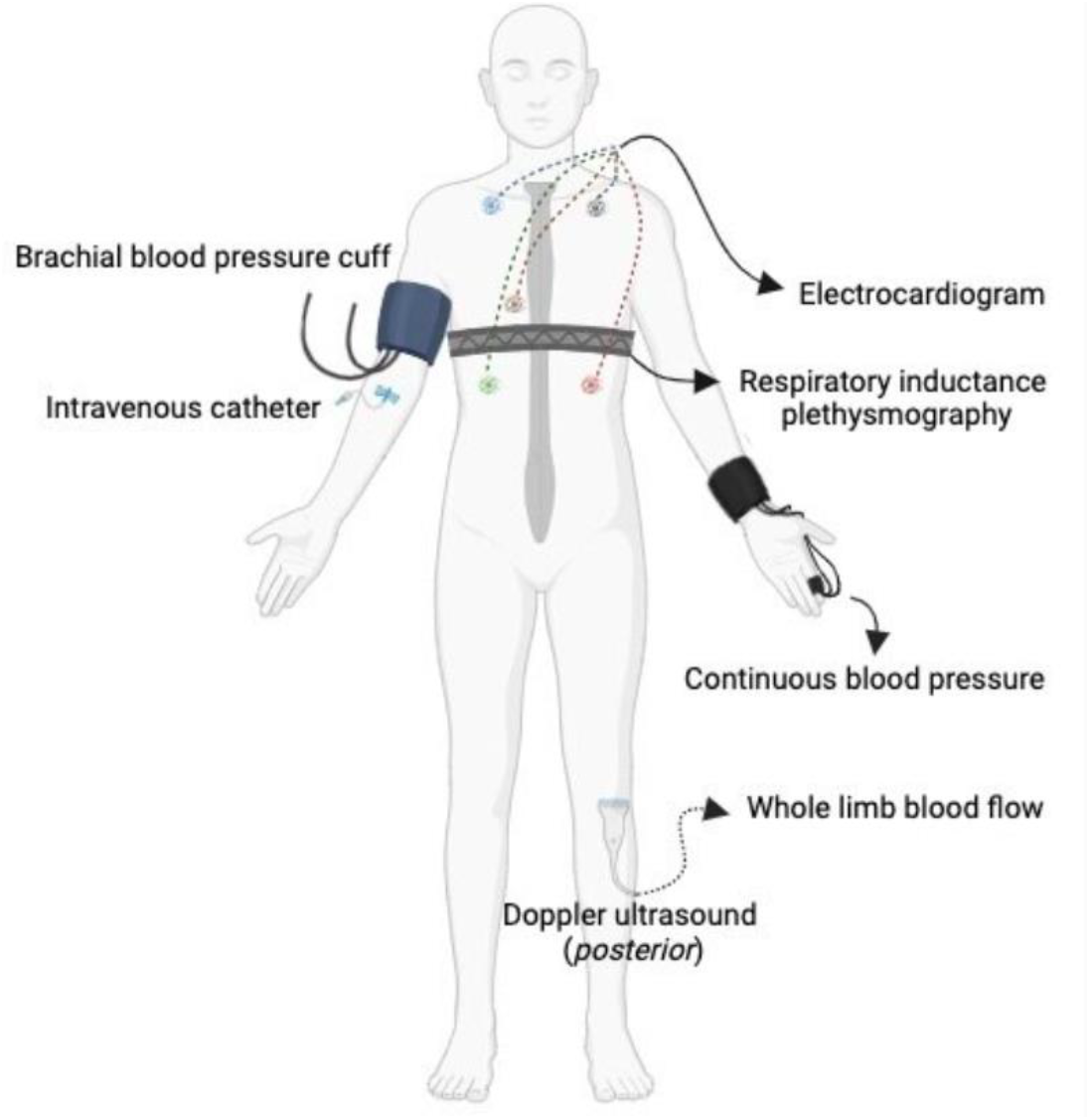
Experimental recording setup for battery of laboratory-based autonomic tests.

### Identifying effects of sympathetic inhibition

With impaired sympathetic regulation through the spinal cord, individuals with “autonomically complete” SCI would be expected to have a lack of sympathetic inhibitory control below the level of injury. To identify the ability to inhibit sympathetic activity, heart rate/blood pressure variability and intravenous bolus phenylephrine responses were recorded. Supine recordings of continuous blood pressure and heart rate were made during five minutes of paced (0.25Hz) breathing.^27^ Data were digitized and stored at 1000 Hz for offline analysis of heart rate and blood pressure variability. Greater values of unbuffered blood pressure changes and increased reliance on compensatory heart rate mechanisms for blood pressure homeostasis would signal impairments in sympathetic inhibition. For boluses of intravenous phenylephrine, the Oxford method was used to pharmacologically increase blood pressure.^28^ Three successive escalating doses (25-175 μg) were given to achieve targeted increases in systolic blood pressure of 15-40 mmHg. At least eight minutes (approximately two half-lives)^29^ were allowed between administration of doses to limit carryover effects. As phenylephrine pharmacologically induces widespread vasoconstriction, compensatory sympathetic inhibition can be quantified through magnitude of change in lower extremity vascular resistance, blood pressure, and heart rate (Table 2).

**Table 2:** Metrics and scientific rationales for each of 18 component analyses encompassing an *autonomic phenotype*. LF = Low Frequency; HF = High Frequency; MAP = Mean Arterial Pressure; AUC =Area Under the Curve; RRi= R-R interval; SBP= Systolic Blood Pressure

| Test | Metric | Rationale |
| --- | --- | --- |
| **Sympathetic Inhibition** |  |  |
| Heart Rate and Blood Pressure Variability | <sup>A</sup> Log transformed LF blood pressure variability | Fluctuations may be low as only able to compensate with changes in heart rate, with driving changes in blood pressure dampened by low vascular tone.<sup>42</sup> |
|  | <sup>B</sup> Log transformed HF heart rate variability | Fluctuations may be decreased given limited or absent afferent signaling from below the level of injury.<sup>43, 44</sup> |
|  | <sup>C</sup> HF blood pressure and heart rate cross spectral phase | Typical feed forward relationship of heart rate into blood pressure may be reversed following SCI given lack of vascular control below the level of injury.<sup>44</sup> |
| Bolus Intravenous Phenylephrine | <sup>D</sup> ΔMAP AUC | Larger AUC represents greater reactant vasoconstriction without ability to actively inhibit baseline sympathetic activity. |
|  | <sup>E</sup> Baroreflex sensitivity | Serves as a metric for vagal outflow to compensate to changes in blood pressure.<sup>35</sup> |
| | <sup>F</sup> Percent $\Delta$ lower extremity vascular resistance | Given known differences in resting vascular resistance below the level of injury,<sup>36</sup> percent change serves as a measure of vessel responsiveness to phenylephrine and ability to inhibit baseline sympathetically-mediated vasoconstriction. |
| **Supralesional Sympathetic Activation** |  |  |
| Hand Cold Pressor | <sup>G</sup> ΔMAP AUC | Decreased AUC when supralesional sympathetic activation is unable to cause vasoconstriction below the level of injury in SCI. |
| | <sup>H</sup> ΔRRi AUC | Quantification of tachycardic pressor response through vagal withdrawal and sympathetic activation. Expected to be normal after SCI and serving as a checkpoint on degree of noxious input. |
| | <sup>I</sup> Percent $\Delta$ lower extremity vascular resistance | Decreased with above level sympathetic activation that is unable to pass through the injured cord to cause vasoconstriction. |
| Valsalva's Maneuver | <sup>J</sup> ΔSBP baseline to phase II nadir | Greater fall in SBP demonstrates relative loss of sympathetically mediated vasoconstriction below the level of injury.<sup>10</sup> |
|  | <sup>K</sup> Pressure Recovery Time | Decreased sympathetic activation is captured as a delay in this metric following expiratory pressure release.<sup>39</sup> |
|  | <sup>L</sup> Valsalva Ratio | With intact vagal mechanisms after SCI, this ratio would be expected to be normal. Inclusion serves as a capture of abnormalities during release of expiratory pressure.<sup>45</sup> |
| **Sublesional Sympathetic Activation** |  |  |
| Foot Cold Pressor | <sup>M</sup> ΔMAP AUC | Quantification of sympathetic activation originating below the level of injury. |
| | <sup>N</sup> ΔRRi AUC | Metric of systemic sympathetic engagement. In the setting of sensory and autonomically complete SCI, increase MAP AUC would not generate tachycardia, but instead bradycardia due to lack of communication through the injured cord and absence of typical pressor response. |
| | <sup>O</sup> ΔMAP AUC * ΔRRi AUC | Capture of the key interplay dynamics between blood pressure change and RRI change. In clear cases of dysreflexic response, large positive values represent increase in blood pressure with lengthened RRI (bradycardia).<sup>12</sup> |
| Bladder Pressor | <sup>P</sup> ΔMAP AUC | As in <sup>M</sup> |
| | <sup>Q</sup> ΔRRi AUC | As in <sup>N</sup> |
| | <sup>R</sup> Percent $\Delta$ lower extremity vascular resistance | Measure of sympathetically mediated vasoconstriction when stimuli originate below the level of injury. |

### Identifying effects of supralesional sympathetic activation

Individuals with autonomically complete SCI impacting sympathetic activation would be further expected to have blunted responses to supralesional sympathetic activation, being able to only recruit sympathetic activity above the level of injury. To characterize this, both a hand cold pressor test and the Valsalva maneuver were performed. Hand cold pressor test, which would be above level stimulus in individuals with neurological level of injury below C7, was begun with three minutes of supine, resting baseline recordings. Individuals then placed their hand in 0° C water for three minutes to provide a measured, mildly noxious stimulus. During the test, participants were asked at one-minute intervals how their pain ranked on a visual analog scale. The hand was then removed and actively rewarmed.^19^ Effects of supralesional sympathetic activation are able to be assessed with the hand cold pressor test though quantifying the pressor response, recording magnitude of change in lower extremity vascular resistance, blood pressure, and heart rate (Table 2). Valsalva maneuver was performed at least three times to assess the ability to rapidly engage whole-body sympathetic-induced vasoconstriction. In individuals with SCI, phase IIb of this response (representing an attempted compensatory tachycardia and systemic vasoconstriction to stabilize blood pressure) is commonly impaired due to inability to recruit sympathetic vasoconstriction below the level of injury.^11^ To complete this maneuver, individuals breathed out against a mouthpiece with a small air leak in place to ensure an open glottis. Expiratory pressure was maintained as close as possible to 30 mmHg for 15 seconds with subsequent quiet recovery.

### Identifying effects of sublesional sympathetic activation

To identify sublesional sympathetic activation, mildly noxious stimuli were initiated below the level of injury. Both a foot cold pressor and bladder pressor test were performed. For the foot cold pressor test, three minutes of supine, resting baseline was first recorded. Individuals then had their foot wrapped in 0° C ice packs for three minutes. Just as with the hand cold pressor test, participants were asked at one-minute intervals how their pain ranked on a visual analog scale. The foot was then actively rewarmed. For the bladder pressor test, individuals with SCI first had a urinary catheter placed to drain their bladder. Following drainage and several minutes to re-equilibrate in a semi-supine position, three minutes of quiet resting baseline were recorded. The bladder was then filled with room temperature sterile water to capacity, where it remained for five minutes to provide a static stretch. Throughout both the filling phase and static stretch phase of this simple cystometry, bladder pressure was maintained between 35-40 cmH_2_0 to apply a consistent afferent stimulus to stretch receptors,^30,31^ regardless of bladder capacity/compliance. If bothersome symptoms or concerns for safety were present, filling was stopped and the bladder was drained. Bladder pressor test was not completed for uninjured controls and normative data was derived from the foot cold pressor for this cohort (see data analysis).

### Data analysis

For heart rate and blood pressure variability, R-R interval and systolic blood pressure time series were analyzed using custom peak detection algorithms (Matlab, Mathworks Inc.). All tracings were visually inspected for artifacts prior to analysis, with subsequent interpolation at 4 Hz and linear detrending. Cross-spectral relationships were calculated using a Welch’s modified periodogram^32^ and three-linear detrended segments were divided into five equal, 50% overlapping portions and smoothed via a Hanning window. Spectral densities were integrated over low (0.05 – 0.15 Hz) and high (0.20 – 0.30 Hz) frequencies to generate average powers. Confidence intervals and precision of estimate for the transfer function were derived based on the level of coherence from standard random process theory.^33^ Cross-spectral phase was weighted by precision to obtain the most accurate means for statistical analysis. In this way, unreliable estimates received appropriately small weights when group averages and statistics were computed.^34^ For bolus intravenous phenylephrine, change in mean arterial pressure from baseline was calculated. Area under the curve from the inflection in blood pressure (usually 12-15 beats after intravenous administration) to two minutes after was calculated and normalized to dose of phenylephrine and individual body weight for drug distribution as a measure of the pressor response. Baroreflex sensitivity from the inflection point in R-R interval to the initial peak was also calculated and included for all values with systolic blood pressure increase of greater than 10 mmHg.^35^ Lower extremity vascular resistance was calculated. As baseline resistance tends to be significantly lower for individuals with SCI,^36^ following confirmation of this assumption, percent change in resistance was also calculated from baseline to a fifteen second window encompassing the initial peak R-R interval. The average value across this window was chosen as opposed to other timepoints of resistance, as at this initial point, compensation from vagally mediated bradycardia (and subsequent impact on blood pressure) is in response to the full, known dose of phenylephrine prior to rapid metabolization.

For the hand cold pressor test, resting mean baseline values of mean arterial pressure (MAP), R-R interval, and lower extremity vascular resistance were calculated. Area under the curve (AUC) for MAP and R-R interval were calculated for the three-minute immersion in ice water as markers of sympathetically mediated vasoconstriction and tachycardia. Vascular resistance was calculated in 30 second windows for the three minutes of immersion, with the window from 60 to 90 seconds compared as percent change to baseline as an additional marker of relative sympathetic recruitment. This window was chosen to allow for maximal vasoconstriction originating from preserved sympathetic outflow without the effect of systemic catecholamines, which may confound results from prolonged stimuli.^37^ Valsalva maneuvers were visually inspected for error and adequate expiratory pressure maintenance, with phases defined per accepted standards.^38^ Baseline values of MAP and R-R interval duration were established from a quiet resting 30 seconds prior to initiation. Nadir of systolic blood pressure during phase IIb of the Valsalva maneuver was compared to baseline as a marker of sympathetic activation. Pressure recovery time, an accepted measure of sympathetic activation,^39^ was calculated as the time from nadir of phase III MAP to return to baseline MAP. Valsalva ratio, calculated as the peak R-R interval for phase IV to nadir in R-R interval for phase III was also computed.^40^

Similar to the hand cold pressor test, AUC for MAP and R-R interval were calculated for the foot cold pressor. In this context, with below level sympathetic activation, while MAP may increase, R-R interval would be expected to increased (i.e. bradycardia) in an individual with SCI and lack of full autonomic regulation. To fully capture this dynamic of increased blood pressure and bradycardia, a third metric, the product of MAP and R-R interval AUC was calculated. This allows for differentiation of a small blood pressure increase with a large, buffered bradycardia in an individual with SCI from a small blood pressure increase with minimal brady or tachycardia in an uninjured control who may have minimal pain associated with the test. The bladder pressor test also had mean baseline values of MAP, R-R interval, and vascular resistance calculated. AUC for MAP and R-R interval were calculated and normalized to the duration of the test (as some individuals had their tests abbreviated due to symptomatic autonomic dysreflexia). Given differences in resting vascular resistance, following validation, percent change in vascular resistance was calculated for individuals with SCI at initial peak MAP as above. As uninjured controls did not complete this bladder pressor testing, normative values for the foot cold pressor test were utilized for comparisons to the SCI cohort.

Key metrics derived for each test appear in Table 2, with accompanying scientific rationale for each. To test each metric’s normality, Shapiro-Wilk statistics were calculated for the control cohort. Non-normal distributions were log transformed when consistent with typical precedence in literature (i.e. heart rate and blood pressure variability).^41^ As a marker of test reproducibility and analytic psychometrics, repeat testing was performed on a subset of both uninjured controls and individuals with SCI, with mean Z-scores compared for repeat tests.

### Data representation

One major barrier to broader comprehension of autonomic testing is the lack of clarity in normative data. To address this, we utilized our uninjured control population to establish such norms for each of the tests in our battery. For each test, three key representational metrics were selected (Table 2). **Admittedly, no single metric from any autonomic test encapsulates the multifaceted interplay at work. It was thus our hope that broad reporting approach aided bringing this to light**. Using this normative data, results for each of the 18 total metrics were then converted to Z-scores and plotted radially, serving as a digestible graphical representation of autonomic regulation customized for individuals with SCI (Figure 2). In this representation, a Z-score of greater than 2.33, eclipsing the 99^th^ percentile of expected results for each metric, is treated as abnormal. Such spikes of abnormality describing the degree of autonomic dysregulation can be rapidly identified using these visualizations, which we refer to as *Autonomic Phenotypes*.

**Figure 2:**
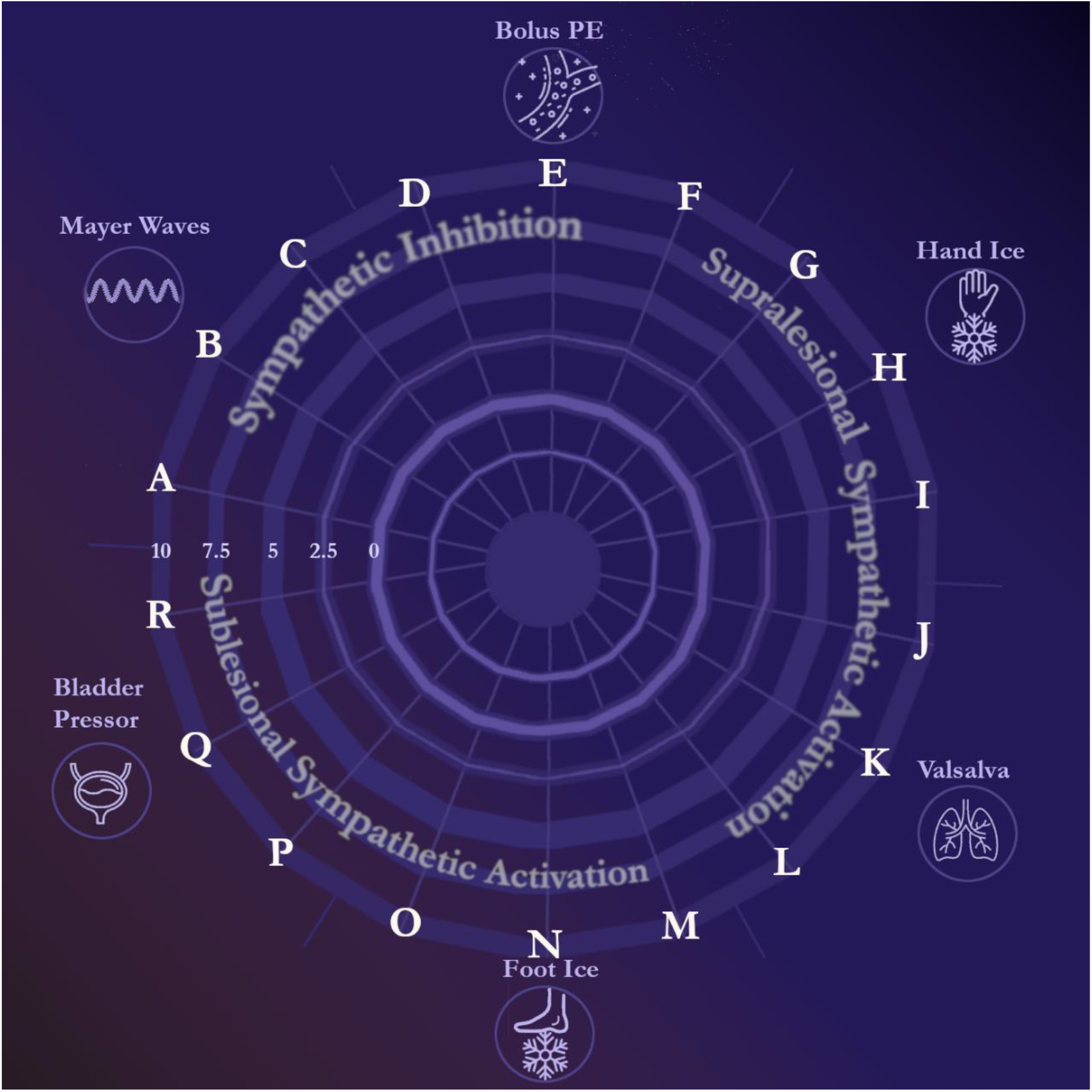
Graphical layout of an *autonomic phenotype*, with letters corresponding to metrics listed in Table 2. Concentric circles outward from zero represent increased Z-scores for each metric, allowing results from the full testing battery to be represented as one shape. Metrics are grouped by laboratory test and tested domain, facilitating a snapshot assessment of how normal (or abnormal) sympathetic inhibition and activation may be.

## Results

Twenty-three uninjured controls contributed normative control data. ADFSCI scores (mean 9.3 ± 11.0, standard deviation) and COMPASS-31 (6.4 ± 7.2) for this cohort demonstrate low levels of baseline autonomically mediated symptoms. As an example of how these tests are able to differentiate autonomic dysfunction after SCI, results for this same battery of tests appear for 11 individuals with SCI. Symptoms of autonomic dysfunction as measured by ADFSCI (58.4 ± 47.0, p=0.006) and COMPASS-31 (32.5 ± 13.4, p<0.001) were both significantly higher in those with SCI. Further demographic characteristics for both cohorts were well matched and appear in Table 1.

Tests to quantify sympathetic inhibition, paced breathing for heart rate and blood pressure variability and bolus intravenous phenylephrine, generally demonstrated normally distributed data with few outliers (Figure 3). Mean log of low frequency blood pressure variability power was similarly between controls (0.3 ± 0.5) and those with SCI (0.4 ± 0.4, p=0.46). Log of high frequency heart rate variability power was higher in controls (3.0 ± 0.5) than those with SCI (2.4 ± 0.7, p=0.02). Mean high frequency phase (n=13 in control cohort with adequate coherence) was 19.5 ± 11.6°, equating to a feed forward mechanism of heart rate into blood pressure with a phase lead of 401 ± 239 ms. This was not significantly different from those with SCI (n=6 with adequate coherence), where the mean values was −19.3 ± 39.7° (phase lag of 399 ± 822 ms, p=0.07). For bolus phenylephrine, MAP AUC normalized to kg of body weight and μg of medication had a mean value of 0.056 ± 0.053 mmHg·s/kg·μg for controls. This showed early differences between uninjured controls and those with SCI, who exhibited significantly greater sensitivity (Figure 3, mean 0.27 ± 0.22 mmHg·s/kg·μg, p=0.03). Mean baroreflex sensitivity was 24.7 ± 9.3 ms/mmHg for the control cohort, which was significantly greater that those with SCI (12.3 ± 6.8 ms/mmHg, p=0.004). Resting baseline vascular resistance was 8.3 ± 4.7 U for controls and 4.7 ± 1.5 U for those with SCI (p=0.003). Given this significant baseline difference, percent change was used. Mean percent increase in lower extremity resistance for 6 ± 11% for a standardized 75 kg person receiving a 75 μg dose for controls and 20 ± 21% for those with SCI (p=0.12, absolute change 0.4 ± 1.0 U for control cohort vs 0.9 ± 0.6 U for SCI cohort, p=0.08).

**Figure 3:**
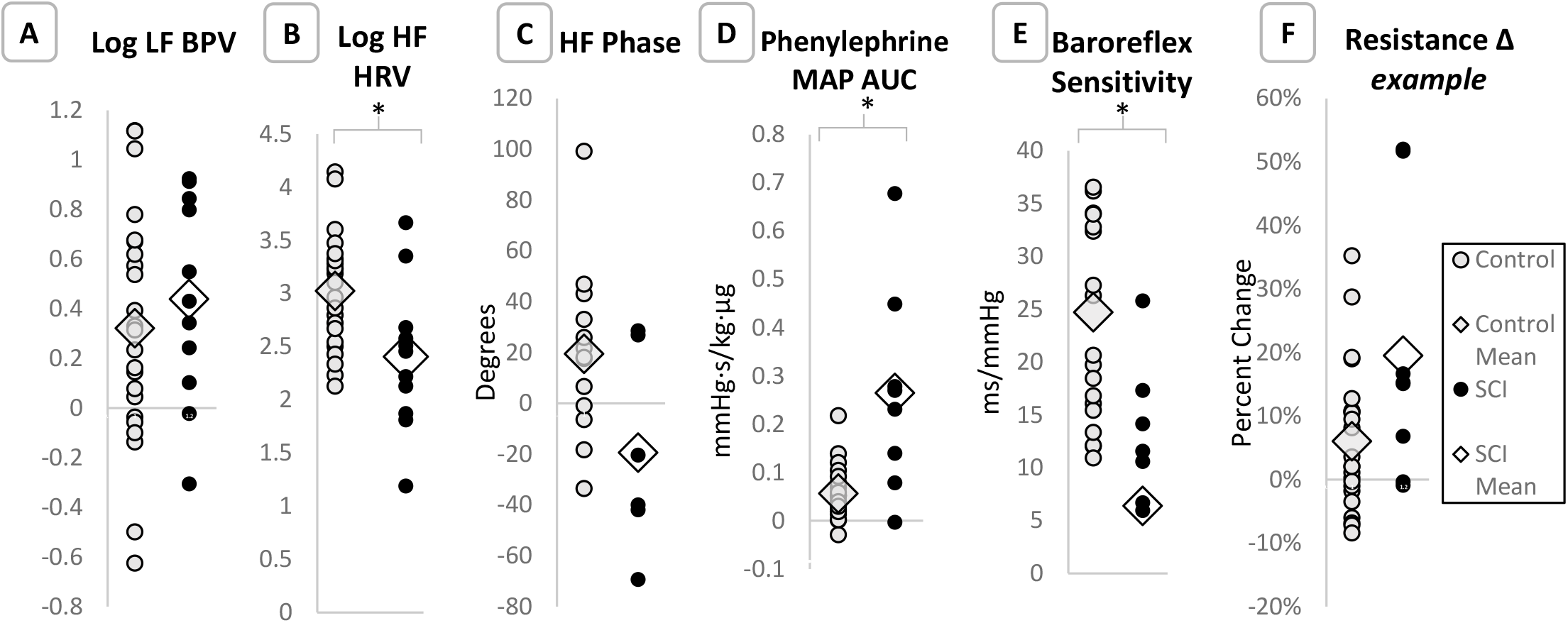
Results of sympathoinhibitory testing. **A)** Log of control LF blood pressure variability, *W*= 0.97, p=0.78; **B)** Log of control HF heart rate variability, *W*= 0.97, p=0.60; **C)** Control HF phase relationship of cross spectrum between heart rate (R-R interval) and (systolic) blood pressure variability, *W*= 0.95, p=0.62; **D)** Control cohort change in MAP normalized to kg of body weight and μg of phenylephrine administered, *W*=0.90, p=0.08; **E)** Control baroreflex sensitivity; *W*= 0.90, p=0.11; **F)** Control percent change in lower extremity vascular resistance set to a standardized 75 kg individual receiving 75 μg of phenylephrine, *W*=0.93, p=0.24. *Indicates statistical significance in comparison between control cohort and SCI cohort preliminary data. LF= Low Frequency; HF= High Frequency; BPV= Blood Pressure Variability; HRV= Heart Rate Variability; MAP= Mean Arterial Pressure; AUC= Area Under the Curve. SCI= Spinal Cord Injury

For tests of the effects of supralesional sympathetic activation, hand cold pressor demonstrated a mean increase in MAP AUC of 3137 ± 3255 mmHg·s in the control cohort, with similar changes in those with SCI (mean AUC 1700 ± 1871 mmHg·s, p=0.23). R-R interval AUC in the control cohort was −18.0 ± 11.8 s^2^, showing a tachycardic response, and was also similar to those with SCI (−15.2 ± 17.1 s^2^, p=0.64, Figure 4). Baseline lower extremity vascular resistance was significantly higher in the control cohort (7.1 ± 3.0 U) than those with SCI (4.3 ± 1.7 U, p<0.001). As such, percent changes in resistance were again calculated. The control cohort increased lower extremity vascular resistance by 33 ± 29% with the hand cold pressor test. This was not statistically different from the small cohort of individuals with SCI (mean increase of 22 ± 21%, p=0.20, absolue change of 2.3 ± 1.4 U for control cohort vs 0.9 ± 0.8 U for SCI cohort). Valsalva maneuver demonstrated a mean phase IIb drop of 2.4 ± 13.3 mmHg in the control cohort, which was significantly less than those with SCI (decrease of 17.4 ± 18.3 mmHg, p<0.001). Mean pressure recovery time was 1.7 ± 1.1 s in controls. By comparison, individuals with SCI exhibited significant delays (mean pressure recovery time of 8.7 ± 6.8 s, p<0.001). Mean Valsalva ratio for the control cohort was 1.88 ± 0.45, similar to those with SCI (mean 1.72 ± 0.50, p=0.16).

**Figure 4:**
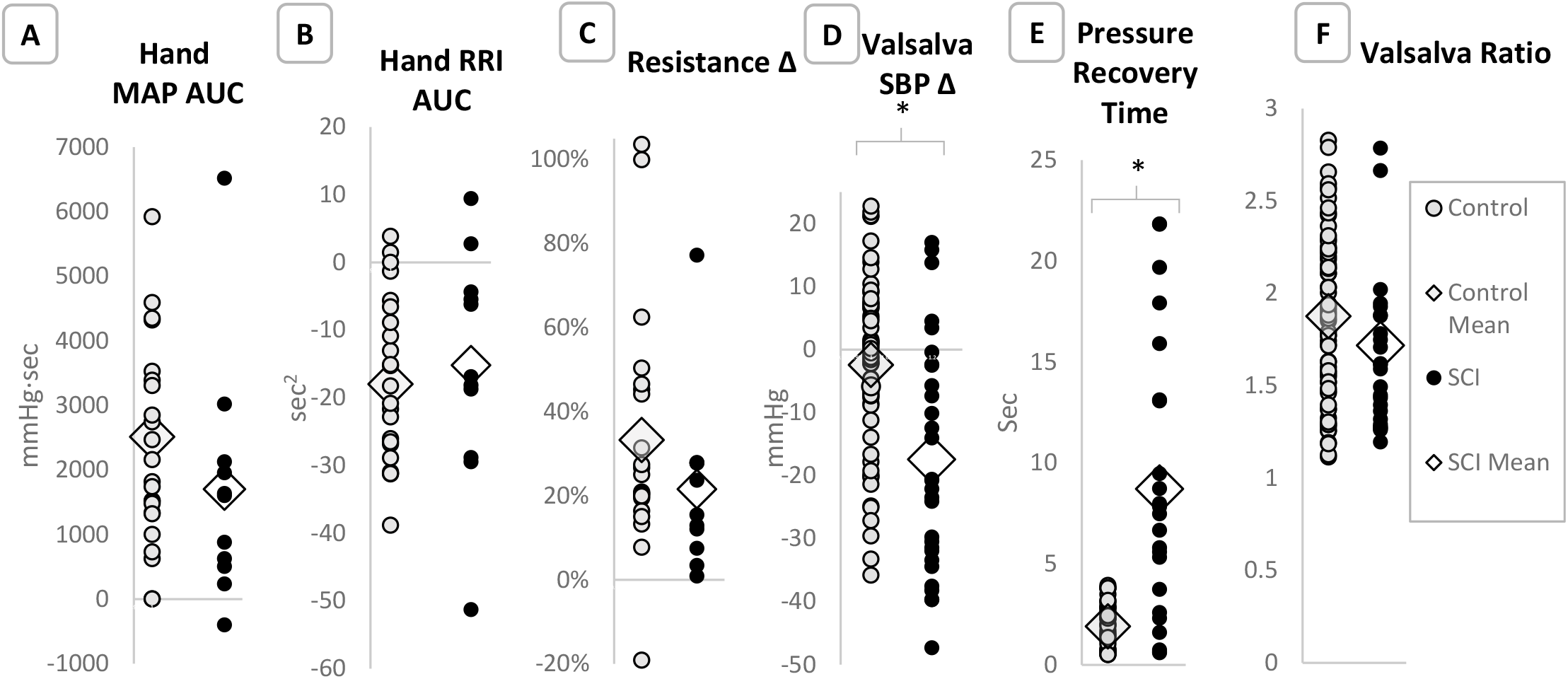
Results of supralesional sympathetic activation testing. **A)** Change in MAP area under the curve during hand cold pressor test for control cohort, *W*=0.616, p<0.001; **B)** Change in R-R interval area under the curve for control cohort, *W*=0.97, p=0.66; **C)** Percent change in control cohort lower extremity vascular resistance with hand cold pressor test, *W*= 0.83, p=0.002.; **D)** Control cohort change in systolic blood pressure from baseline to nadir of phase IIb during the Valsalva maneuver, *W*=0.97, p=0.10; **E)** Control cohort pressure recovery time during the Valsalva maneuver; *W*= 0.90, p<0.001; **F)** Control cohort Valsalva Ratio, *W*=0.97, p=0.08. *Indicates statistical significance in comparison between control cohort and SCI cohort preliminary data. MAP= Mean Arterial Pressure; AUC= Area Under the Curve; RRi= R-R wave interval; SBP= Systolic Blood Pressure

For tests centered on the effects of sublesional sympathetic activation, foot cold pressor resulted in a mean MAP AUC increase of 826 ± 1004 mmHg·s for the control cohort. These results were similar for individuals with SCI (521 ± 1013 mmHg·s, p=0.42). R-R interval AUC decreased 4.5 ± 8.2s^2^ in the control cohort compared to an increase of 5.1 ± 10.0 s^2^ (p=0.01, Figure 5), representing a bradycardic response for those with SCI. The composite product of these results, meant to differentiate a normal pressor response of hypertension and tachycardia from a dysreflexic response of hypertension with compensatory bradycardia, demonstrated a mean value of −1655 ± 3397 mmHg·s^3^ in controls and 1333 ± 2794 mmHg·s^3^ in those with SCI (p=0.04). Bladder pressor test resulted in mean MAP AUC increase of 1377 ± 1674 mmHg·s (normalized to five minutes of testing) for controls and 5790 ± 7239 mmHg·s for those with SCI (p=0.13). Similarly normalized R-R interval AUC decreased 7.6 ± 13.7 s^2^ for the control cohort. This was significantly different in the SCI cohort, with a mean increase of 40.3 ± 29.5 s^2^ (p=0.01). Lower extremity vascular resistance increased 26 ± 17% from baseline during testing for controls, and 43 ± 63% for those with SCI (p=0.48).

**Figure 5:**
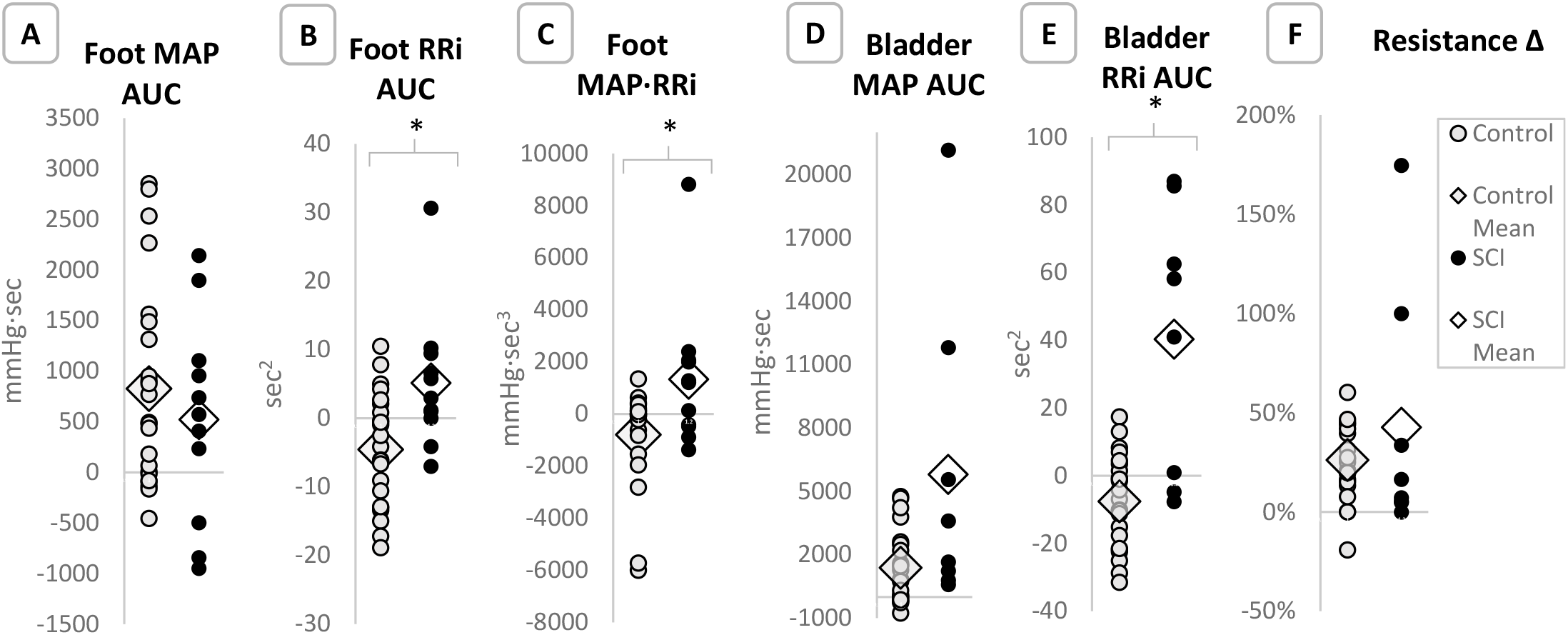
Results of sublesional sympathetic activation testing. **A)** Control cohort change in MAP area under the curve during foot cold pressor test, *W*=0.89, p=0.02; **B)** Change in R-R interval area under the curve for control cohort during foot cold pressor test, *W*=0.97, p=0.78; **C)** Control cohort product of MAP and RRi area under the curve to capture interplay dynamics, *W*= 0.69, p<0.001; **D)** Control cohort change in MAP area under the curve during bladder pressor test, normalized to 5-minute duration, *W*=0.89, p=0.02; **E)** Change in R-R interval area under the curve during bladder pressor test for control cohort; *W*= 0.97, p=0.78; **F)** Percent change in lower extremity vascular resistance during bladder pressor test, *W*=0.87, p=0.007. *Indicates statistical significance in comparison between control cohort and SCI cohort preliminary data. MAP= Mean Arterial Pressure; AUC= Area Under the Curve;

As a global data representation of cardiovascular autonomic testing results, an example autonomic phenotype plot is presented for an uninjured control (Figure 6A) and three individuals from the SCI cohort (Figure 6B-D). While the example autonomic phenotype for the uninjured control individual visually appears quite round, discrete spikes of abnormality are present for the example individuals with SCI. Notably the individual in Figure 6B, with a C7 AIS A injury, demonstrates global cardiovascular autonomic impairments in sympathetic inhibition and supra/sublesional sympathetic activation. The individual in Figure 6C, with a T8 AIS A injury, has clear deficits in sympathoinhibition and some difficulty with supralesional sympathetic activation, though has normal response to sublesional sympathetic activation. The individual in Figure 6D, with T10 AIS A SCI, has near normal cardiovascular autonomic regulation, with only moderate impairments in supralesional sympathetic activation.

**Figure 6:**
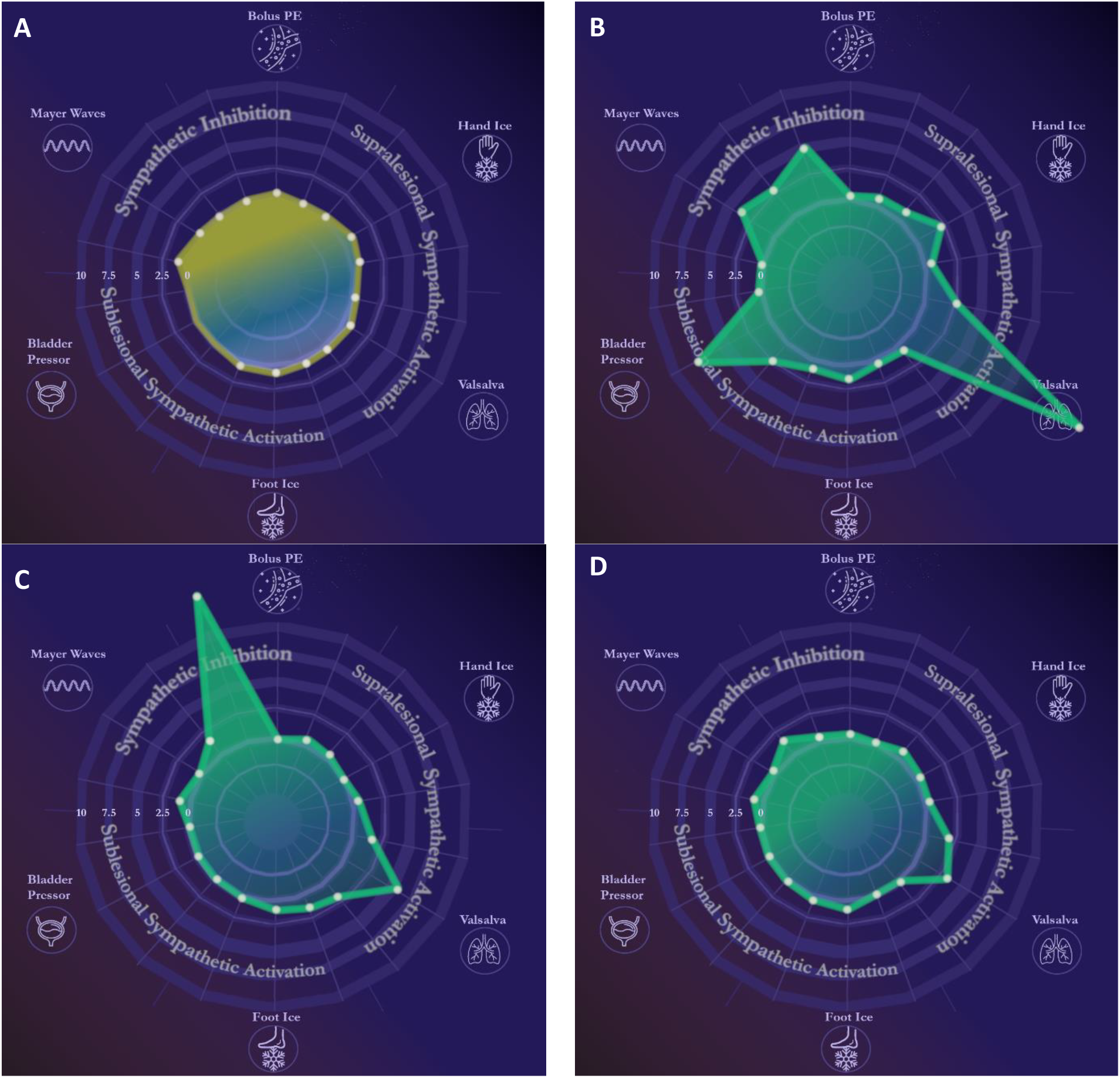
Examples of *autonomic phenotypes* for an uninjured control **(A)** an individual with C7 AIS A SCI demonstrating global cardiovascular autonomic dysfunction **(B)**, an individual with T8 AIS A SCI demonstrating impairments in sympathetic inhibition and supralesional sympathetic activation **(C)**, and an individual with T10 AIS A SCI and only mild impairments in supralesional sympathetic activation **(D)**.

For data reproducibility and test psychometrics, two uninjured controls and three individuals with SCI were retested a mean of 422 days after initial baseline testing (range 127 – 996 days). Baseline and repeat mean Z-scores appear in Figure 7. Average mean Z-scores for controls were 0.79 at baseline and 0.80 at follow up. The mean change over this time period on an individual basis was 0.15 ± 0.01. For the three individuals with SCI, average mean Z-scores was 1.57 at baseline and 1.52 at follow up. The mean change in Z-score between tests was 0.19 ± 0.04. In aggregate, mean Z-scores were unchanged in this mixed group of individuals on repeat testing (p=0.77).

**Figure 7:**
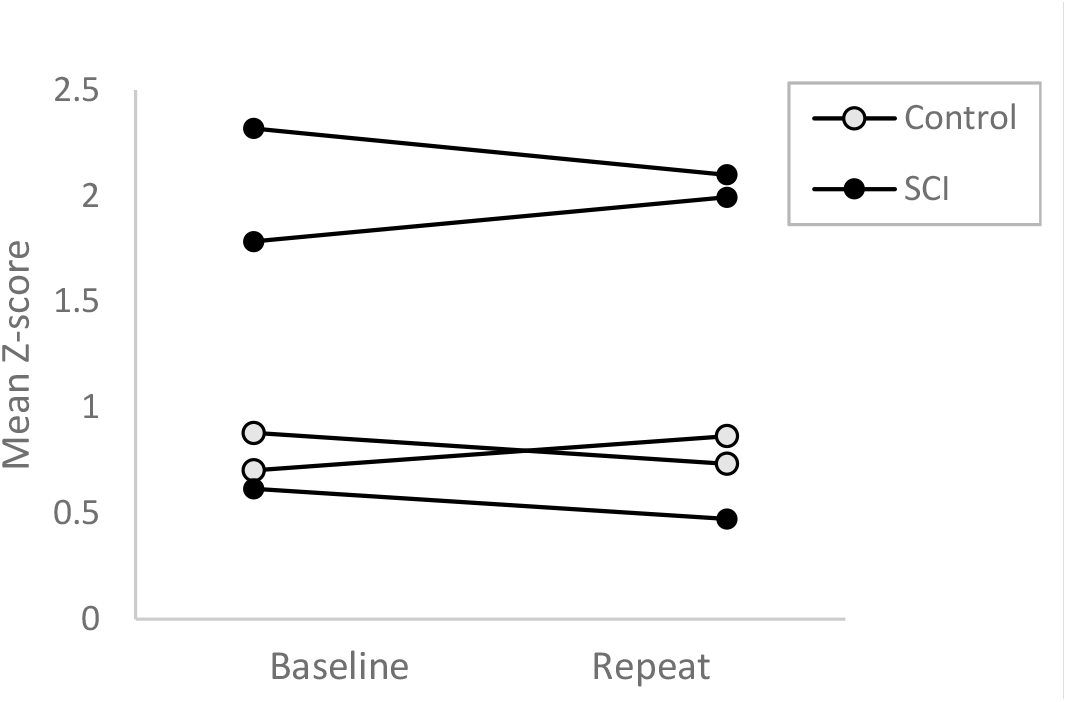
Mean Z-scores across testing battery at baseline and repeat testing.

## Discussion

Results of the uninjured controls cohort and the small sample of individuals with SCI demonstrates unique insights using this battery of strategically assembled cardiovascular autonomic tests. Establishing these normal uninjured control values is vitally important to perform as an element of this testing battery, as it allows identification of abnormal responses after SCI. Additionally, tests which demonstrated similar responses, such as heart rate response during hand cold pressor, serve as important checkpoints of normality. Even in this small preliminary sample of individuals with SCI, clear differences in the domains of sympathetic inhibition and supra/sublesional sympathetic activation were demonstrated. Further, early results demonstrate strong reproducibility with minimal, non-significant changes in mean Z-scores on repeat testing. Individuals with SCI and more baseline autonomic dysfunction did have a greater change in this mean Z-score on repeat testing. This may represent changes in autonomic function/recovery or variance in the context of different magnitudes of resting afferent stimuli (i.e. subacute changes in bowel or bladder patterns).^46^ As there is limited literature on autonomic recovery after SCI, further investigation is needed.

Notably, the field of SCI Medicine has long acknowledged the importance of assessing autonomic function^47^ and moving beyond the current clinical model of deferring to bedside motor/sensory assessments.^48^ The international standards to document remaining autonomic function (ISAFSCI) have existed since 2012,^49^ though largely gathered clinical reports of conditions such as resting bradycardia or hypotension. Correspondingly, recent surveys have noted that only 10% of physicians utilize these standards and many confuse them for a standardized dataset as opposed to an assessment of autonomic function.^50^ The ISAFSCI was subsequently updated in 2021,^51^ with new objective measures of temperature, forced vital capacity, and heart rate and blood pressure in supine and seated positions. This updated ISAFSCI nobly attempts to implement laboratory autonomic measures for individuals with SCI within the clinic using available resources. However, it has yet to be seen if these new limited measures will better assess autonomic function or how some procedures, such as repeated ischemia from auscultatory brachial blood pressure at one-minute intervals, may confound findings.^52^

Beyond the ISAFSCI, Berger *et al*. previously did test an objective group of laboratory assessments for autonomic function after SCI.^5^ Their battery similarly utilizes Valsalva’s maneuver, though alternatively tests heart rate response to deep breathing, heart rate and blood pressure responses to a 45° head-up-tilt test, and sympathetic skin responses at the wrist and ankle. While an important advancement for the field, these tests focus on supralesional sympathetic activation, with minimal attention to sympathetic inhibition or activation below the level of injury. This produces an incomplete representation of autonomic regulation for individuals with SCI.

Evolving past a single metric of autonomic regulatory dysfunction, our graphical autonomic phenotypes provide a more nuanced presentation of cardiovascular autonomic regulation-one which we hope is more beneficial in defining an autonomically complete SCI. Defining an autonomically complete SCI is critical before discriminative statistics on testing batteries such as ours can be applied. These statistical approaches, in larger sample sizes, will refine the key metrics which need to be tested to meet this definition and which recordings provide little additional benefit. It was our intention in our testing battery to cast a wide net and include a range of translationally focused cardiovascular autonomic tests, though it is very possible that other established tests or new tests may provide improved or more clinically accessible predictive metrics.

Even as a small group, individuals with SCI demonstrated significant differences in eight of the 18 metrics from our testing battery. This encompassed tests of sympathoinhibition, supralesional sympathetic activation, and sublesional sympathetic activation. Notably for sympathoinhibition, responses to bolus phenylephrine were significantly larger. This is likely due, in part, to vessel hyperreactivity from α1-adrenoreceptor upregulation^53^ in combination with the impaired compensatory baroreflex sensitivity (Figure 3 and demonstrated previously^13^). For supralesional sympathetic activation, few differences were noted for the hand cold pressor test. This is not unexpected as a mild, subjective experience for an uninjured control may generate minimum pressor response in a similar way to someone with SCI who is unable to recruit sympathetic activation below their level of injury. The Valsalva maneuver, by comparison, demonstrated broad impairments in rapid sympathoexcitation, as has previously been shown.^10^ For sublesional sympathetic activation, both foot and bladder pressor tests demonstrated significant differences between controls and those with SCI. Notably, there was a wide distribution of responses for those with SCI, with several of the more pronounced hypertensive/bradycardic events (Figure 5 B-F) occurring when these stimuli elicited mild autonomic dysreflexia. Our testing metric of the product of hypertensive response with bradycardia demonstrates promise at capturing this unique dynamic, though further testing is needed to differentiate if this provides added value.

Interestingly, applying our testing battery in the exemplar three individuals with SCI, clear patterns of autonomic (dys)regulation appear. In the individual with C7 AIS A injury (Figure 6B), testing revealed a greatly reduced capacity to activate their sympathetic nervous system from above the level of injury. This may place them at increased risk for orthostatic hypotension and other clinical scenarios requiring rapid increase in blood pressure. This individual has similar, significant dysfunction in their activation of sympathetic activity below the level of injury which is compounded by the inability to inhibit sympathetic activity. This likely clinically translates to a high risk of autonomic dysreflexia (from exaggerated sublesional sympathetic activation) with more pronounced hypertension (given lack of sympathoinhibitory ability).

In the individual with T8 AIS A SCI (Figure 6C), classic clinical teaching would suggest that they are at low risk for autonomic dysfunction (with autonomic dysreflexia typically taught to be associated with neurological levels of injury at or above T6 and even temperature dysregulation associated with injuries above T8). However, upon detailed laboratory autonomic testing, this individual has noted impairments in cardiovascular sympathetic inhibition and moderate difficulty with sympathetic activation through their SCI. Contrasting this, the individual with T10 AIS A SCI (Figure 6D) appears largely normal, with only one individual Z-score falling slightly outside of the expected variability. This is, in itself, is surprising, as despite having a motor and sensory complete SCI (AIS A), this individual is able to alter not only heart rate and systemic blood pressure, but lower extremity vascular resistance within normal limits. This demonstrates that despite being motor/sensory complete, this individual has evidence of what may be considered an autonomically incomplete SCI.

Our goal in this study has been to rigorously quantify and characterize autonomic dysfunction in individuals with SCI. We’ve felt this step was necessary prior to defining autonomically complete SCI, enabling discriminating key testing metrics for eventual clinical translation. As many of these tightly controlled laboratory testing conditions (only morning evaluations, pre-test fasting, extensive testing resource needs, etc.) are less replicable in clinical practice, key metrics will need to be validated in this setting. More broadly, while application of this work is targeted for assessment of cardiovascular autonomic regulation in individuals with SCI, this testing and visualization construct may be further adapted to other disease processes where there are less discrete locations of neurologic damage.

### Limitations

While introducing a novel combination of autonomic tests and graphical visualization, our control dataset (n=23) is currently smaller than ideal, as evidenced by non-normality in some expected metrics. Enrollments are ongoing to grow this dataset and produce more stable metrics over time. Yet even in this smaller sample, clear differences emerge in individuals with SCI compared to controls. In multiple cases, these differences exceed 10 standard deviations from the mean and demonstrate clear dysfunction in a manner not previously visualized. Further, while we included six tests of autonomic function, there exist other well-established tests such as tilt test and handgrip which could provide additional insights. As these tests both assess supralesional sympathetic activation and come with unique challenges in the SCI population (i.e. straps for the tilt table introducing potential below level pain, potential lack of volitional muscle control in upper extremities to perform handgrip), they were excluded from this current testing battery.

## Conclusions

This novel combination of established laboratory autonomic tests provides a strong foundation for assessing autonomic regulation after spinal cord injury. Our *autonomic phenotype* graphical representation allows for a rapid snapshot of this data to aid in understanding of autonomic deficits and consensus on what constitutes and autonomically complete SCI. Collectively, this represents an important advancement in our understanding of autonomic dysfunction after spinal cord injury.

## Data Availability

All data produced in the study will be made available following publication and subsequent analyses upon reasonable request to the authors.

## Acknowledgements

The primary data for this study is funded by NIH/NICHD (K23HD102663), with further contributions from the National Institutes of Health National Center of Neuromodulation for Rehabilitation, the National Center for Complementary and Integrative Health, the National Institute on Deafness and Other Communication Disorders, and the National Institute of Neurological Disorders and Stroke. NIH/NICHD Grant Number P2CHD086844 which was awarded to the Medical University of South Carolina. Pilot work for this study was funded through the Foundation for Physical Medicine & Rehabilitation under the Milbank Award. The contents are solely the responsibility of the authors and do not necessarily represent the official views of the funders.

